# A pilot pre-post trial with and without subsidy to promote safe backyard poultry-raising practices to prevent exposure to poultry and poultry feces in rural Bangladesh

**DOI:** 10.1101/2024.11.12.24317180

**Authors:** Laura H. Kwong, Jesmin Sultana, Elizabeth D. Thomas, Mohammad Rofi Uddin, Shifat Khan, Ireen Sultana Shanta, Nadia Ali Rimi, Md. Mahbubur Rahman, Peter J. Winch, Tarique Md. Nurul Huda

## Abstract

**Background:** Backyard poultry-raising is common in rural Bangladeshi households. Raising poultry contributes to fecal contamination of the domestic environment, increasing children’s exposure to enteric pathogens, including *Campylobacter*, which has been associated with child stunting.

**Objective:** To investigate the effectiveness of a behavior change communication and counseling intervention to encourage households to confine poultry outside of the household dwelling in a shed at night and improve poultry feces management.

**Methods:** We conducted a two-arm pre-post pilot study. Households in both arms received the behavior change communication and counseling intervention. Households in the subsidy arm also received ~23 USD for the construction of a poultry shed for nighttime housing. We administered a household survey and spot-check before and after intervention implementation among 37 subsidy and 42 non-subsidy households.

**Results:** At endline, 58% of all households had a poultry shed (87% of subsidy and 33% of non-subsidy households) and the percentage of households confining all poultry outside the house the previous night was significantly higher at endline (33%) compared to baseline (2.5%) (prevalence difference [PD]: 30 percentage points [pp]; 95% CI: [19, 41]). Additionally, more households had no visible poultry feces piles inside the house compared to baseline (PD: 26pp 95% CI: [12, 41]), but there were no significant differences in the number of poultry feces piles in the courtyard or veranda.

**Discussion:** Our intervention effectively encouraged households to confine poultry outside of household dwellings at night and to maintain an indoor living space free of poultry feces. Households were willing and able to construct a shed even without a subsidy. Households that received a subsidy were more likely to construct a shed. Future studies should assess if housing all poultry outside the household dwelling reduces children’s exposure to poultry feces enough to mitigate health risks associated with poultry ownership.

## Introduction

Backyard poultry raising is common across the globe (Birhanu et al. 2023). In Bangladesh, 80% of rural households raise backyard poultry, where poultry is the primary source of animal protein for households (Dolberg 2007) and income for women (Sultana et al. 2012). In this setting, humans live near poultry: in one study among poultry-raising households, 98% reported poultry scavenging in the yard, 93% said poultry roam freely inside homes during the day, and 37% reported children touching, carrying, or playing with poultry in the past two weeks (Shanta et al. 2017). Many households keep poultry inside at night and confine poultry only intermittently during the day (Passarelli et al. 2021; Lowe et al. 2022; Headey and Hirvonen 2016).

Close contact with poultry may have public health implications. Children can be exposed to zoonotic pathogens through contact with poultry or poultry feces (Matilla et al. 2018; Penakalapati et al. 2017; Headey et al. 2017). Findings from observational studies suggest that free-roaming poultry in the domestic environment and housing poultry inside the household dwelling at night have been associated with poor child health outcomes such as stunting and environmental enteropathy (Headey and Hirvonen 2016; Kaur, Graham, and Eisenberg 2017; George et al. 2015). A cross-sectional study analyzing demographic and health surveys from 30 countries found that ownership of livestock and poultry increased the risk of all-cause mortality (Kaur, Graham, and Eisenberg 2017; Zambrano et al. 2014).

Close contact with poultry and poultry feces increases the risk of *Campylobacter*, *Salmonella* (Zambrano et al. 2014), *Cryptosporidium* infections (Moore et al. 2016), as well as avian influenza (Sultana et al. 2012). *Campylobacter* infections, caused primarily by consumption of contaminated water and unpasteurized milk, exposure to poultry and wild birds, and person-to-person transmission, are one of the most common bacterial causes of gastroenteritis among children worldwide (Kaakoush et al. 2015). In low-income countries, *Campylobacter* is endemic; the multi-country MAL-ED study estimated that 85% of children had been infected by *Campylobacter* in their first year of life (Amour et al. 2016) and that *Campylobacter* was responsible for 12.1 episodes of diarrhea per 100 child-years, despite *Campylobacter* infections often being asymptomatic (Platts-Mills et al. 2018). In the Global Enteric Multicenter Study, *Campylobacter* was a leading pathogen associated with moderate-to-severe diarrhea among children <5 years old in South Asia (Kotloff et al. 2013). *Campylobacter* spp. infections have been associated with poor linear growth, increased intestinal permeability, increased intestinal and systemic inflammation, and Guillan-Barre Syndrome (Amour et al. 2016; Nyati and Nyati 2013). *Campylobacter* infections are more common among children living with poultry compared to children not living with animals (Zambrano et al. 2014). In Egypt, children from households with one or more poultry positive for *Campylobacter jejuni* were found to have 3.9 times higher odds of being infected with *C. jejuni* compared to those without any positive birds (El-Tras et al. 2015). These studies suggest that confining poultry to separate them from children could improve child health.

Confining poultry in improved poultry sheds could also improve poultry health compared to non-confinement or typical in-home confinement arrangements. There is no single definition for an improved poultry shed, but generally they are defined as being outside, having multiple compartments, and natural ventilation (Lambrecht, Waid, et al. 2023). Housing poultry of different types and ages to avoid competition over food and reduces stress that can cause pecking out other birds feathers and cannibalism (Food and Agriculture Organization 2004). Adequate ventilation improves air quality inside the shed by reducing the concentration of particulate matter, pathogens, and gases such as ammonia, all of which can harm poultry health and diminish productivity (Li et al. 2020; Al-Kerwi et al. 2022). Ventilation can prevent the shed from becoming too hot or humid, which can easily cause death among poultry due to their lack of sweat glands and compensate for high animal density (Weaver 2002). There are several ways that elevating a poultry shed off the ground can benefit poultry health (Food and Agriculture Organization 2004). Firstly, elevation can protect poultry (and their eggs and feed) from snakes and burrowing predators such as rodents. An elevated floor can also prevent water from runoff or flooding or high moisture from clay soil from entering the shed and causing the growth of pathogens or mold in bedding or the floor of the coops; moist coops can also wet feathers and are also more likely to cause frostbite in the winter. Air can circulate under an elevated coop, allowing for cooler temperatures in the summer. In settings where poultry are confined in fenced-in runs when not inside the coop, the area under the coop can add to the size of the run and function as an area of the run that is shaded and protected from the rain. Some improved sheds also provide a perch, as perches allow birds to separate from each other, reducing stress and aggression and improving welfare and production efficiency (Bist et al. 2023).

Given the potential health risks associated with poultry, interventions encouraging poultry production may generate greater net health benefits if participants also receive training in hygienic poultry management practices, particularly the reduction of children’s physical exposure to poultry and their feces. However, there is limited research on interventions to prevent exposure to poultry and poultry feces. Little is known about the types of confinement structures and behaviors that could effectively separate children from poultry and poultry feces and are feasible and acceptable in low-income settings (Oberhelman et al. 2006). As such, we aimed to develop a poultry and poultry feces management intervention to separate young children from poultry feces in backyard poultry-raising households in rural Bangladesh and understand the influence of these interventions on young children’s exposure to poultry and poultry feces.

## Materials and Methods

### Setting

We conducted the study in the Fulbaria sub-district of Mymensingh district in north-central Bangladesh. We selected Mymensingh based on its proximity to Dhaka, and prior findings that approximately half of the poultry-raising households in this area house poultry inside their household dwelling at night.

Fulbaria is typical of rural sub-districts in Bangladesh; it contains 13 unions, each with several villages. In each village, there are compounds of two to eight related households, clustered around a central courtyard but the poultry are typically owned by the households.

### Study Design

We conducted a two-phase study to explore how to reduce children’s exposure to poultry and poultry feces in rural Bangladesh. In Phase I, we conducted formative research to identify and refine existing local strategies that could separate children from poultry feces and develop new technologies. The formative research, which included transect walks across all districts in Bangladesh, qualitative interviews, and trial of improved practices, are described in a forthcoming manuscript. Based on the findings of Phase I, we then designed a neighborhood-based behavior change and counseling intervention to promote nighttime confinement of poultry outside the house and improve poultry feces management. In Phase II, we conducted a two-arm pilot pre-post trial to investigate the effectiveness of this intervention, with and without monetary support, to encourage backyard poultry-raising households to 1) confine all poultry outside the household living space at night, specifically by keeping them overnight in an improved poultry shed outdoors; 2) wash hands with soap and water after contact with poultry, poultry products, and poultry feces; 3) remove poultry feces immediately; and 4) dispose of poultry feces in a specific pit. Households in both arms received the behavior change communication and counseling intervention. One arm additionally received a monetary subsidy of 2000 BDT (USD ~23) for the labor and/or material costs associated with constructing a poultry shed. The subsidy arm allowed us to assess households’ need for financial support to build a poultry shed.

### Selection of study households and participants

We compared the socio-demographic characteristics (e.g., literacy rate, household ownership, population density, male-to-female ratio, sanitation coverage, safe drinking water provision at the household level, and electricity supply) of 13 unions in the Fulbaria sub-district and excluded six because they were urban, hilly, or outliers in terms of the aforementioned characteristics. From the seven eligible unions, we used a random number generator to randomly select two unions; unions were considered as the unit of randomization because they are large enough to reduce the likelihood that participants in the non-subsidy union would learn about the subsidy offered in the subsidy union. From each selected union, we selected one village based on convenience, poultry raising practices, and distance from the Fulbaria sub-district center. In each village, trained enumerators identified the village’s center and then went door-to-door in a clockwise direction from the center to identify compounds in which 1) each of the households had at least one adult chicken (at least two months old), 2) at least one household housed poultry inside their dwelling at night, and 3) at least one household had a child 6-59 months old. Enumerators visited 13-21 compounds, totaling 35-62 households, to list 30-38 eligible households per village. They prepared a hand-drawn map with landmarks of the village, the relative position of the compounds, the number of households in each compound, and the presence of poultry and children in those households. Using the maps, the study team selected neighboring compounds to each other to create a supportive environment for adopting recommended behaviors and creating new poultry-raising norms. We excluded compounds with households that engaged in commercial poultry farming (50+ birds) and compounds with a single household not adjacent to other compounds. In each village, we enrolled 7-8 compounds with a total of 19-21 households. The primary poultry-raiser was defined as the household member who owned and cared for the poultry and was the primary decision-maker for the poultry. In rural Bangladesh, primary poultry-raisers are most often female (Shanta et al. 2017). Study investigators used a random number generator to randomly allocate each village to the subsidy or non-subsidy intervention arm. The household enrollment process was completed in February 2020.

### Sample size

We enrolled 80 households in this pilot study. Prior pilot studies in similar settings suggest that the inclusion of 50 households per arm allows for exploring trends in behavioral uptake (Desai et al. 2015; Oberhelman et al. 2006). Due to budgetary constraints, we were limited to 40 households per arm.

### Masking

Intervention delivery and data collection were conducted by separate teams. Data collection staff were not told whether the village had been assigned to the subsidy or non-subsidy arm.

### Intervention design and delivery

Based on the formative research findings, we developed an intervention approach called Neighborhood-based Environmental Assessment and Planning (NEAP). NEAP is a novel participatory approach that incorporates principles of household-based assessments and an ecological model approach to behavioral determinants, considering contextual, psychosocial, and technological factors at multiple levels (societal, community, household, individual and habitual) likely to influence behavioral outcomes in the development of intervention content and delivery (Dreibelbis et al. 2013; Thomas 2021). We also took guidance from research on self-assessment of health hazards in the domestic environment, which can be a helpful tool for both identifying hazards and possible solutions (Tomita et al. 2014; Morgan et al. 2005). Intervention activities included a community engagement meeting, training carpenters on the construction of improved sheds, group meetings and household visits with the primary poultry raisers, and engagement meetings with male household members (Table 1). This pilot study promoted four key behavioral recommendations: 1) confine all poultry out of the household dwelling at night; 2) wash hands with soap and water after contact with poultry, poultry products, or poultry feces; 3) remove poultry feces as soon as you see them; and 4) dispose of poultry feces in a specific site away from children’s reach.

**Table 1:** Characteristics of the study population at baseline (February 2020)

| Indicators | Non-subsidy arm (n = 42)<br>% or Mean ± SD | Subsidy arm (n = 38)<br>% or Mean ± SD | Total (n = 80)<br>% or Mean ± SD |
| --- | --- | --- | --- |
| Status of the respondent |  |  |  |
| Female | 100% | 100% | 100% |
| Has child 6-59 months of age | 50% | 58% | 54% |
| Average number of individuals in the household | 4.0 ± 1.4 | 4.3 ± 1.4 | 4.1 ± 1.4 |
| Average number of households in the compound | 3.9 ± 1.6 | 3.0 ± 1.1 | 3.5 ± 1.4 |
| Survey respondent's mean age (years) | 37 ± 14 | 35 ± 12 | 36 ± 13 |
| Survey respondent's formal education (years) |  |  |  |
| No formal education | 43% | 42% | 43% |
| 1-5 years | 36% | 34% | 35% |
| 6-10 years | 19% | 24% | 21% |
| More than 10 years | 2.4% | 0% | 1.3% |
| Primary occupation of the main earning member of the household (top 6) |  |  |  |
| Day/unskilled laborer (domestic, agricultural, or migrant work within the country) | 33% | 26% | 30% |
| Work on own farm or as a sharecropper | 24% | 24% | 24% |
| Rickshaw/Van puller/Boat driver | 21% | 5.3% | 14% |
| Skilled worker other than carpenter/carpenter (long term contracted laborer) | 0% | 18% | 8.8% |
| Business owner | 9.5% | 7.9% | 8.8% |
| Carpenter/carpenter | 2.4% | 7.9% | 5.0% |
| Has access to an improved latrine<sup>a</sup> | 67% | 55% | 61% |
| Household assets |  |  |  |
| Has electricity (including solar) | 100% | 95% | 98% |
| Television (functional) | 4.8% | 21% | 13% |
| Refrigerator (functional) | 2.4% | 7.9% | 5.0% |
| Mobile phone | 88% | 95% | 91% |
| Smartphone | 36% | 16% | 26% |
| Household poultry ownership |  |  |  |
| Chickens (any age) | 100% | 100% | 100% |
| Average number of chickens (any age) per household | 8.8 ± 7.0 | 8.5 ± 7.9 | 8.6 ± 7.4 |
| Healthy<sup>b</sup> adult chickens (>2 months age) | 86% | 92% | 89% |
| Sick<sup>c</sup> adult chickens (>2 months age) | 0% | 5.3% | 2.5% |
| Egg-laying chickens | 50% | 55% | 53% |
| Healthy chicks (<2 months age) | 64% | 50% | 58% |
| Sick chicks (<2 months age) | 14% | 11% | 13% |
| Ducks (any age) | 21% | 63% | 41% |
| Average number of ducks (any age) per household | 0.48 ± 1.1 | 3.0 ± 3.8 | 1.7 ± 3.0 |
| Household ruminant ownership |  |  |  |
| Pigeons | 0% | 11% | 5% |
| Bulls/milk cows/buffaloes | 26% | 61% | 43% |
| Goats/sheep | 29% | 34% | 31% |

<sup>a</sup> Improved toilet according to JMP: Flush or pour-flush to - piped sewer system, septic tank, pit toilet, ventilated improved pit (VIP) toilet, pit toilet with slab, composting toilet, pit latrine with slab, no bucket and/or hanging toilet
<sup>b</sup> Self-reported by the respondent: Poultry (adult chicken/chick/adult duck/duckling) with no sign symptom of a disease
<sup>c</sup> Self-reported by the respondent: Poultry (adult chicken/chick/adult duck/duckling) with sign symptom of a disease (e.g., fever, convulsion, pox, diarrhea, vomiting)

Each behavioral recommendation was introduced over the course of six group meetings with the primary poultry raisers. We developed three illustrative posters for each behavioral recommendation that served as visual aids during group meetings (Figure S1). The first poster depicted illustrations of common behaviors with negative consequences. In the second poster, photographs of target behaviors with a positive outcome were shown. Prior research has found that in rural Bangladesh, participants responded more positively to illustrations of negative behaviors and photographs of recommended behaviors (Hossain et al. 2020). The final poster presented an enabling technology to facilitate the adoption of target behaviors. The enabling technologies were an improved poultry shed (which we define as an outdoor shed that has multiple compartments, allows for cross-ventilation, and is elevated from the ground), a handwashing station (a bucket with lid and tap, and basin to catch water), a soapy water bottle (a plastic bottle filled with water mixed with detergent powder), and a hoe or spade (for poultry feces removal). A scale model of the improved shed was presented during the meetings. In addition to the behavior-specific posters, an overview poster with illustrations of all four key behavioral recommendations was provided to households at the first group meeting (Figure 1). We asked participants to sign the poster and display them in their homes as a sign of their commitment to the behaviors. Finally, to track their progress toward each recommended behavior, participants were provided with a progress book with illustrations or photographs of small doable actions to take to achieve each behavior.

**Figure 1:**
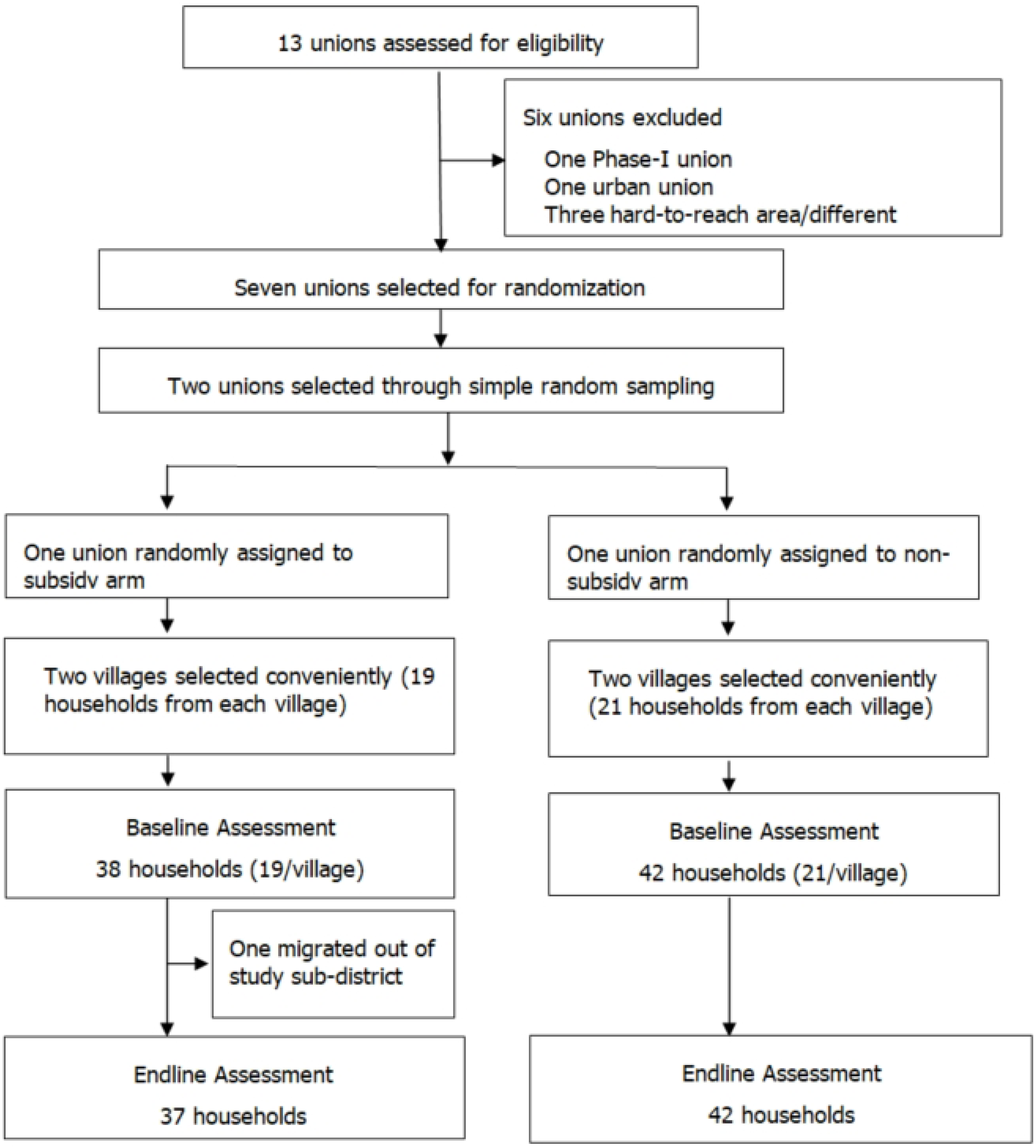
Flow chart of study household selection, enrollment, and participation

**Figure 2:**
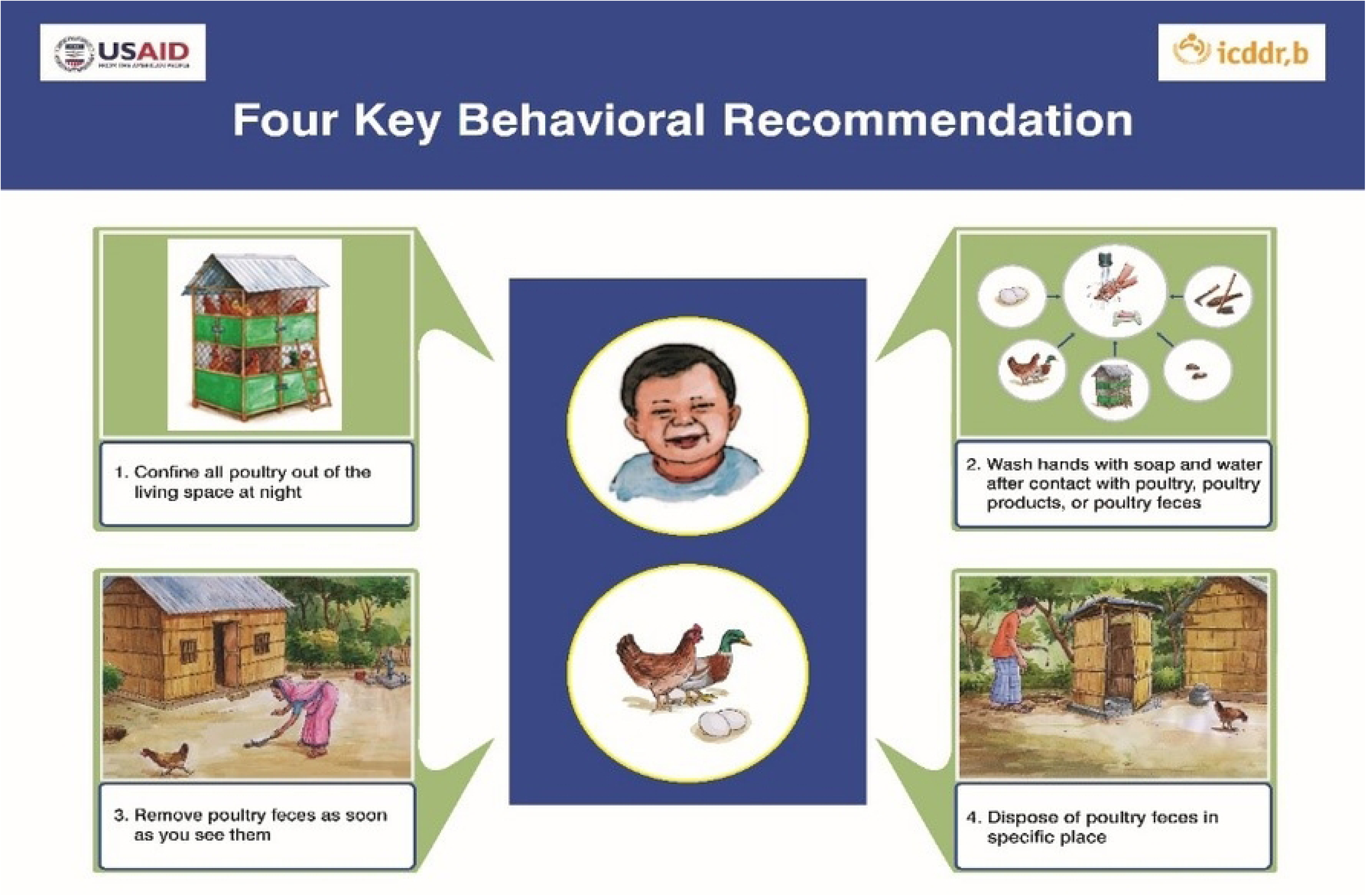
Poster displaying recommended behaviors, shown and discussed at group meetings

We recruited community hygiene promoters (CHPs) (four women and two men) from the study site to facilitate group meetings and household visits. CHPs were >18 years old and had at least eight years of formal education. CHPs received rigorous training and were supervised by project staff. We also trained local carpenters from each study village to introduce them to the project, explain the characteristics of an improved shed, and describe the anticipated role of carpenters in the project.

In each of the two villages, we separated study households into three groups (6-8 primary poultry raisers each). CHPs led sessions for each group every other week and called or visited each participant the day before the meeting to remind them to attend. CHPs conducted group meetings in a convenient place for the households (e.g., courtyard, off-compound location, living room, or veranda). Following the group meeting, they conducted individual household visits in each study household. During household visits, CHPs recorded the household’s progress and challenges toward adopting the previous group meeting’s recommended behaviors, in a separate pictorial progress book, similar to those given to participants. During household visits, CHPs also shared possible solutions to any difficulties reported. Female CHPs led all group meetings and household visits with poultry raisers, all of whom were women.

Male CHPs conducted two engagement meetings with male members of the study households. During these meetings, CHPs provided an overview of the project’s behavioral recommendations, highlighted the important contributions of poultry-raising to the household, and emphasized the critical role that men play in achieving the goal of having both healthy children and healthy poultry.

Due to the COVID-19 pandemic, we delayed implementation of intervention activities until September 2020. At outdoor group meetings, CHPs and participants wore face masks, and individuals stayed 1 m from each other whenever possible. Before each meeting, both as part of the intervention and to follow COVID-19 handwashing recommendations, CHPs and participants washed their hands using the promoted handwashing station and soapy water bottle.

### Data collection

#### Primary and secondary outcomes

The primary outcome of this study was the percentage of households confining poultry outside the household dwelling at night. We defined the household dwelling as the structure where household members usually sleep. The secondary outcomes of the study were the number of poultry feces in the household dwelling and courtyard, access to and/or ownership of an improved night shed, ownership of a specific pit or trash pile for disposing of poultry feces, access to a handwashing station with soap and water, and self-reported handwashing behavior with soap.

#### Quantitative assessment

We conducted a baseline and endline quantitative assessment in all households. The baseline assessment was completed after enrollment in February 2020. The endline quantitative assessment was conducted from January to February 2021, after three months of intervention implementation (mid-September to mid-December 2020). During baseline and endline quantitative assessments, a team of two enumerators (one male and one female) not involved in intervention delivery conducted spot-checks followed by a survey in all study households. The enumerators recorded the data with the CommCare platform on tablet computers.

#### Spot-check

Trained enumerators visited different locations in the household and compound (e.g., household dwelling, courtyard, veranda, and other places) to record signs of poultry (e.g., feathers) and to count visible poultry feces piles. Enumerators counted each pile of poultry feces in each location up to 25 piles and recorded a categorical “>25 piles” if more than 25 piles were present. Enumerators also recorded the presence of different types of poultry housing (e.g., shed, shelter, bamboo cage, corral, and other confinement strategies). We define a “shed” as a poultry confinement structure that is house-like with features that include walls, roof, and door (Thomas 2021). The structure may or may not be independent of other structures, but it must either be a) on the veranda, in the courtyard, or off the compound, or b) transportable to the veranda, courtyard, or off the compound. A “shelter” is a poultry confinement structure that involves built or put-together components that may or may not be joined together. “Simple shelters” cannot be moved outside the house and may have walls, a door, or a roof, but these are not required (Thomas 2021). A bamboo cage is a movable structure placed over poultry to confine them temporarily. Structural elements (e.g., windows) and signs of use (e.g., poultry feces piles) were recorded. Enumerators also observed handwashing stations, poultry feces disposal sites, and the presence of free-roaming poultry in the households. We recorded the availability of water, soap or handwashing agents, and pour devices at the handwashing station.

#### Survey

Enumerators administered a structured survey to the primary poultry-raiser in each enrolled household. The survey included questions on demographics and socioeconomic status (baseline only), poultry-raising practices (day and nighttime poultry confinement, the involvement of other household members in poultry-raising, and decision-making related to poultry and poultry products), poultry health, and children’s observed exposure to poultry and poultry feces. We also asked about psychosocial factors likely to influence poultry-raising practices (e.g., risk perceptions and social cohesion), and information on preferences for poultry housing and available resources and anticipated challenges for building improved poultry sheds.

#### Process monitoring and documentation

We documented intervention activities throughout implementation (Table S1). For each group meeting and household visit, CHPs recorded meeting attendance, duration, and challenges related to intervention delivery in a semi-structured daily record form. Also, icddr,b supervisors observed at least one fortnightly group meeting and two household visits facilitated by each CHP to assess performance, recording their observations in the supervisor monitoring form.

### Data analysis

We summarized socio-demographic characteristics and animal ownership across the intervention arms at baseline. Although villages were randomized to receive the intervention with or without monetary support, there were only two villages per arm, so we analyzed the study with difference-in-difference analysis. To assess the impact of the intervention, we assessed the change in primary and secondary outcomes from baseline to endline for both study arms combined. We also compared the changes in primary and secondary outcomes from baseline to endline between non-subsidy and subsidy arms. We conducted all analyses according to the randomized intervention arm at enrollment (intention-to-treat) without considering session attendance. We also analyzed results considering whether or not the household had a poultry shed at the endline to assess the association between ownership of a poultry shed and poultry management practices. Analysis was conducted using Stata-13.

### Ethical consideration

The protocol (PR-18087) for this study was approved by the Institutional Review Board of icddr,b; the IRB concluded that approval for research involving animals was not required. The enumerators collected informed written consent from the primary poultry-raiser in the study household at enrollment. This trial did not assess any health outcomes and was therefore not registered as a clinical trial.

## Results

In February 2020, the study team enrolled 80 households from four villages of two unions in the Fulbaria sub-district of Mymensingh district. At the endline visit (January-February 2021), one household had migrated out of the study area, resulting in endline data collection for 79 households (Figure 1). At baseline, most characteristics of primary poultry-raisers and their households were similar across the two arms (Table 1). All the primary poultry raisers were female. In the non-subsidy arm, access to an improved latrine and ownership of a smartphone was more common, and ownership of bulls/milk cows/buffaloes, a television, and a mobile phone were less common, than in the subsidy arm.

Seventy percent (n=56) of primary poultry raisers attended all six meetings, and 93% (n=74) attended at least four meetings. At the male engagement meetings, attendance at the first meeting was 89% in the subsidy arm and 85% in the non-subsidy arm but dropped to 76% in the subsidy arm and 39% in the non-subsidy arm for the second male meeting. The intervention did not result in any adverse or unintended effects in either group.

### Confinement of poultry outside at night

At baseline, 2.5% of households reported confining all of their poultry outside their living space at night. An unimproved shed was present in 17% of households and no household had an improved shed. The intervention increased the percentage of households confining all of their poultry outside at night, access to any shed, and access to an improved shed. The percentage of households that reported confining all poultry outside the house the previous night was significantly higher at endline compared to baseline (prevalence difference (PD): 30 percentage points [pp] [19, 41]) (Table 2). The increase occurred in both study arms (non-subsidy arm PD: 25pp [11, 38]; subsidy arm PD: 36 pp [19, 54]) (Table 3). The difference between arms was not statistically significant (DID: 12pp [-10, 34]) (Table 3). The percentage of households that had any type of poultry shed (improved or not) was also higher at endline than baseline (PD: 50pp [40, 62]); most of the new sheds were improved sheds (PD: 58pp [47, 69]) (Table 2). While the increase in access to improved sheds occurred in both study arms (non-subsidy arm PD: 33pp [19, 48]; subsidy arm PD: 87pp [75, 98]), the increase was higher in this subsidy arm (DID: 53pp [35, 72]) (Table 3). Among the households with any poultry shed, 48% of households confined all of their poultry outside the house at night compared to 0% among those who did not have access to a shed (PD: 48pp [28, 68]); among households with access to improved poultry shed, 53% of households confined all of their poultry outside the house at night compared to 3% those without an improved shed (PD: 50 pp [32, 69]) (Table 4).

**Table 2:** Impacts of the Neighborhood-based Environmental Assessment and Planning intervention to reduce the prevalence of poultry sleeping indoors at night and reduce the presence of poultry fecal matter in the household environment.

| Indicators | Baseline<br>(n = 79)<br>% (n) | Endline<br>(n = 79)<br>% (n) | Prevalence<br>difference (baseline<br>to endline)<br>pp (95% CI) |
| --- | --- | --- | --- |
| Percent of households reported confined all poultry outside the living space at night | n = 79<br>2.5% | n = 77<br>33% | **30pp (19, 41)** |
| Poultry housing |  |  |  |
| Percent of households that had any shed | 17% | 67% | **50pp (40, 62)** |
| Percent of households with access to an improved<sup>a</sup> shed located in courtyard | 0 | 58% (46) | **58pp (47, 69)** |
| Percent of households with ownership of an improved shed<sup>b</sup> located in courtyard | 0 | 54% (43) | **54pp (43, 66)** |
| Percent of households has access to handwashing station with water and soap/soapy water | 35% | 46% | 10pp (-4.7, 25) |
| Percent of primary poultry raisers reported washing hands with soap and water after the following events | n = 78 | n = 79 |  |
| After defecation | 77% | 66% | -10pp (-23, 2.4) |
| After cleaning child feces | 9.0% | 11% | 2.5pp (-6.1, 11) |
| Before eating | 21% | 48% | **28pp (14, 42)** |
| Before serving food | 0% | 3.8% | 3.8pp (-0.44, 8.0) |
| Before preparing food | 5.1% | 60% | **54pp (43, 66)** |
| After handling poultry feces | 14% | 65% | **51pp (37, 64)** |
| After feeding poultry/ handling poultry or poultry products | 1.3% | 44% | **43pp (32, 55)** |
| After handling other animal feces | 19% | 34% | **15pp (4.5, 26)** |
| Uncontained poultry feces piles observed inside the household dwelling | n = 79 | n = 78 |  |
| No feces pile | 54% | 81% | **26pp (12, 41)** |
| 1-25 feces piles | 42% | 17% | **-25pp (-40, -11)** |
| >25 feces piles | 3.8% | 2.7% | -1.2pp (-6.8, 4.3) |
| Uncontained poultry feces piles observed in the veranda | n = 60 | n = 61 |  |
| No feces pile | 33% | 39% | 6.0pp (-11, 23) |
| 1-25 feces piles | 58% | 54% | -4.2pp (-21, 13) |
| >25 feces pile | 8.3% | 6.6% | -1.8pp (-5.1, 1.5) |
| Uncontained poultry feces piles observed in the courtyard<sup>c</sup> | n = 79 | n = 77 |  |
| No feces pile | 0 | 1.3% | 1.3pp (-1.2, 3.8) |
| 1-25 feces piles | 15% | 13% | -2.2pp (-12, 7.9) |
| >25 feces pile | 85% | 86% | 0.90pp (-9.5, 11) |
| Poultry feces disposal sites (spot-check confirmed) |  |  |  |
| Specific place | 20% | 65% | **44pp (31, 58)** |
| Used as a fertilizer in crop field or garden | 47% | 20% | **-27pp (-39, -15)** |
| Bush or jungle | 35% | 6.3% | **-29pp (-41, -17)** |
| Drain or ditch | 18% | 3.8% | -14pp (-23, -4.8) |
| Area beyond the courtyard | 1.3% | 7.6% | 6.3pp (-0.13, 13) |
| Water bodies (pond/lake) | 0% | 1.3% | 1.3pp (-1.2, 3.8) |

<sup>a</sup> An outdoor, multi-compartment poultry night-shed with cross-ventilation that is elevated off the ground
<sup>b</sup> Three households had access to an improved shed which were owned by other households of the same compound
<sup>c</sup> During endline, in two households of the non-subsidy arm, the enumerators could not access the courtyard to observe and count the poultry feces

**Table 3:**
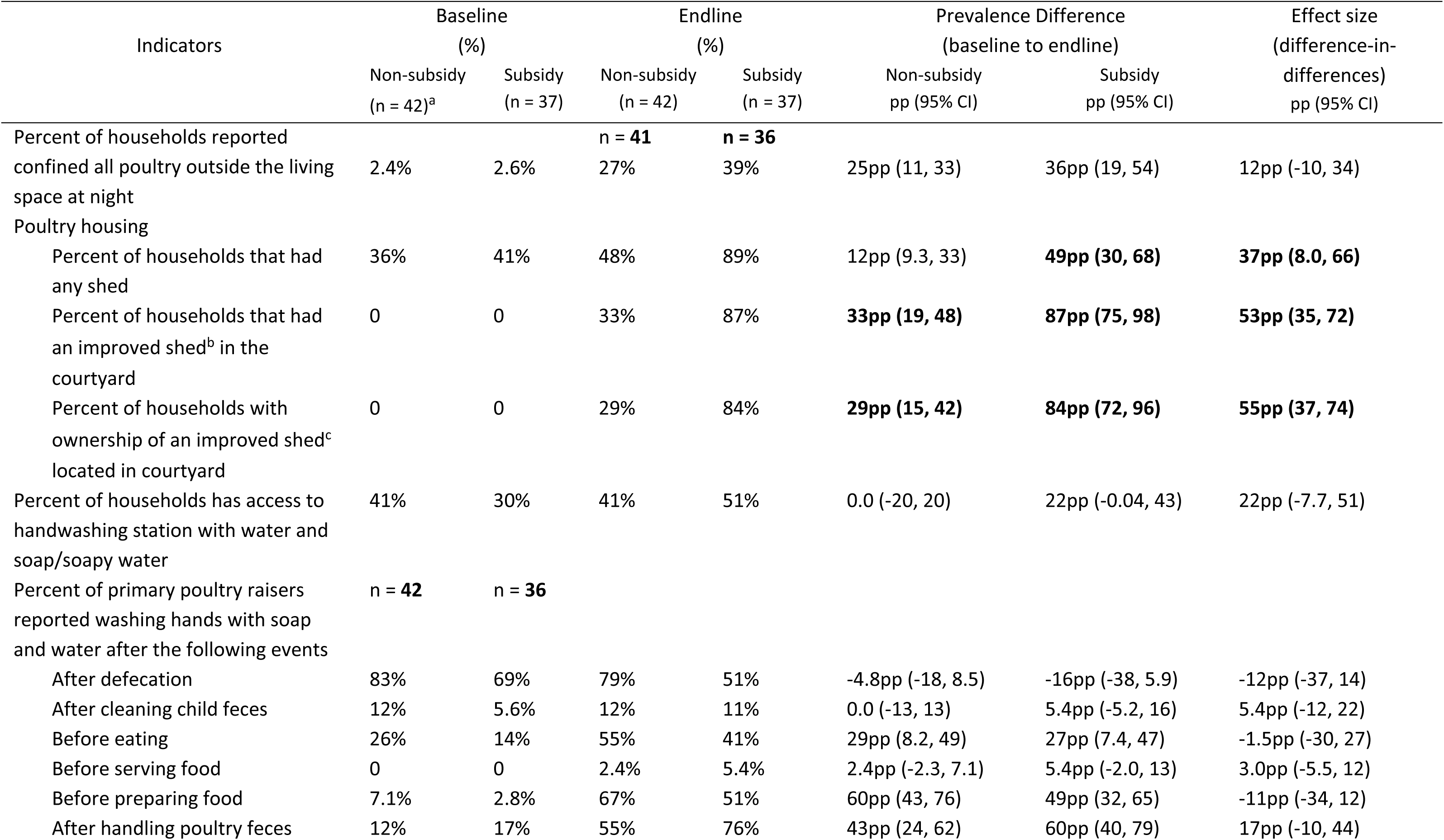
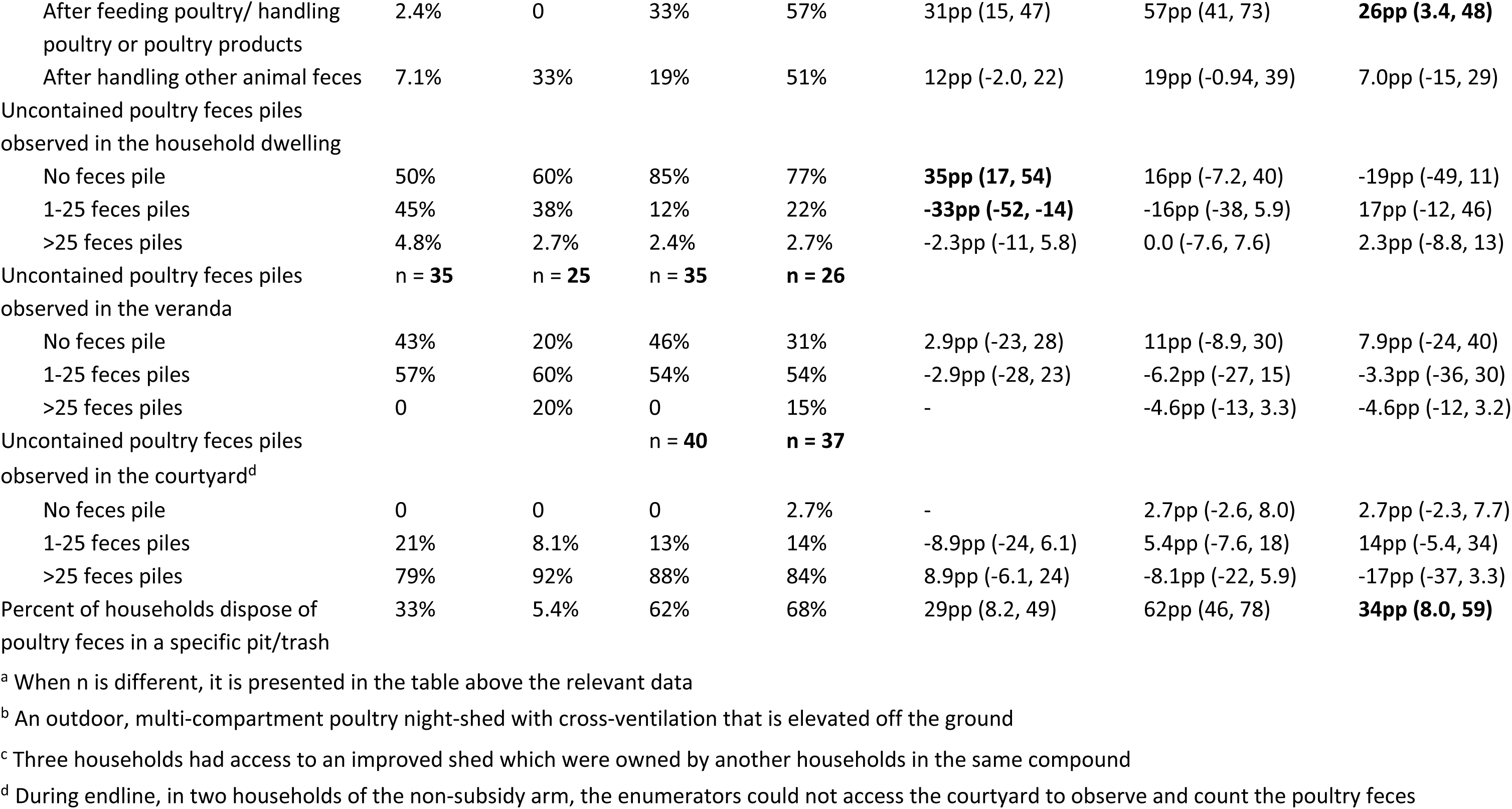
Impacts of the Neighborhood-based Environmental Assessment and Planning intervention to reduce the prevalence of poultry sleeping indoors at night and reduce the presence of poultry fecal matter in the household environment, by study arm.

The 47% of households that had an improved poultry shed but did not confine all of their poultry outside the house at night stated that they were concerned about predators (18/47, 38%) and theft (23/47, 49%). The types of poultry they kept inside were egg laying hens (n=11), adult chickens (n=9), chicks (n=8), ducklings (n=6), and adult ducks (n=3).

Sheds were an average of 15 steps (SD: 7) from the front door of the household. Among 25 households that had an improved poultry shed and children under 5, nine of those reported seeing their children entering into the shed at least once within the previous month and another nine reported observing children entering the shed at least once within the past two months.

### Handwashing

Most primary poultry raisers (77%) reported handwashing with soap after defecation, but handwashing before eating, after handling poultry feces, and after handling other animal feces was practiced by about only one-fifth of primary poultry raisers (Table 2). Fewer than 10% of respondents said they washed hands with soap after cleaning child feces, after feeding poultry / handling poultry or poultry products, or before preparing or serving food.

The intervention did not impact the presence of handwashing infrastructure but did increase handwashing with soap at some key times. There was no statistically significant difference in the percentage of study households observed to have access to a handwashing station with soap and water at baseline and endline (PD: 10pp [-4.7, 25]), but the increase among subsidy households was marginally significant (PD: 22pp [-0.04, 43]) (Table 2; Table 3). The percentage of primary poultry-raisers that reported washing their hands with soap after handling poultry or other animal feces was less than 20% at baseline and increased at endline to 65% after handling poultry feces (PD: 51pp [37, 64]) and 34% after handling other animal feces (PD:15pp [4.5, 26]) (Table 2). Reported handwashing after feeding poultry/ handling poultry or poultry products also increased from less than <5% to nearly 50% (PD: 43pp [32, 55]) and before preparing food (PD: 54pp [43, 66]). Reported handwashing before eating was 21% at baseline and 48% at endline (PD: 28pp [14, 42]). The percentage of primary poultry-raisers who reported washing their hands after feeding/handling poultry or poultry products increased more in the subsidy arm than in the non-subsidy arm (DID: 26pp [3, 48]) (Table 3).

### Poultry feces presence and disposal

At baseline, just over half of households had no uncontained feces (feces outside of the poultry confinement structure) observed inside the house, while 42% had 1-25 piles of feces and the remainder had >25 piles. Verandas were more contaminated, with 58% having 1-25 piles of feces and 8% having >25 piles of feces at baseline. No courtyards were observed without feces: 15% had 1-25 piles of feces and 86% had >25 piles of uncontained feces (Table 2).

The percentage of households that had no uncontained feces inside the household dwelling increased from 54% at baseline to 81% at the endline (PD: 26pp [12, 41]) (Table 2). However, the presence of uncontained feces in the courtyard and veranda remained the same.

At baseline, 47% of households used poultry feces as a fertilizer while 35% threw them in the bush, 18% threw them in a drain or ditch, and 20% disposed of them in a specific place such as a trash pile. The percentage of households that disposed of poultry feces in a trash pile or pit increased substantially after the intervention (PD: 44pp [31, 58%]) (Table 2). The increase was higher among subsidy vs. non-subsidy households (DID: 34pp [8, 59%]) (Table 3).

## Discussion

Based on extensive formative research, we developed and pilot tested an intervention to encourage backyard poultry-raising households to confine poultry outside of the household dwelling in an improved shed at night and improve poultry feces management practices. The Neighborhood-based Environmental Assessment and Planning (NEAP) approach used for this intervention was successful at encouraging poultry-raising households to build (improved and unimproved) poultry sheds, confine poultry outside of the household dwelling at night, and maintain an indoor living space free of poultry feces. Both households that did and did not receive a monetary subsidy constructed improved sheds. The effectiveness of the NEAP intervention may stem from its social mobilization component, which has also been effective at changing latrine construction and open defecation behaviors in Community-Led Total Sanitation (CLTS) programs (Venkataramanan et al. 2018; Pickering et al. 2015).

### Construction and use of improved poultry sheds

At baseline, housing poultry in a shed overnight was uncommon, with only one-fifth of households having sheds and only 2.5% confining all poultry outside at night. The intervention increased the percentage of households with a poultry shed by a notable 50 percentage points. Factors that may have facilitated this high uptake could have been neighborhood commitment and check-ins with study staff to help brainstorm solutions to problems at part of NEAP, training on shed construction provided to local carpenters, sessions for men that emphasized the importance of their engagement for child and poultry health, and the subsidy that some households received. Few other studies have encouraged households to construct their own sheds. One study in Ethiopia required farmers to agree to incurring the cost of providing the 25 chicks they received through the intervention with a “night shelter, daytime enclosure or partitions”. Nine and 18 months after the intervention, “enclosed coop” ownership was higher in the intervention arms than the control arm by 15 percentage points (Passarelli et al. 2021). In addition to the promise made when accepting the intervention, households may have been motivated by needing space for the 25 chickens they received and could not accommodate within their house or calculating that an enclosed coop would provide a high return on investment if it protected the high-value chickens. Another study, in rural Bangladesh, implemented a homestead food production intervention in which one component was providing households with partial reimbursement for the cost of a constructed, improved poultry shed (Wendt et al. 2019; Lambrecht, Waid, et al. 2023). Post intervention, 77% of women in the intervention group owned a shed compared to 28% in the control group. In the intervention group, 68% of all women owned an improved shed and among poultry owners 75% used the shed for poultry.

Even when households have a shed, they may not use it to confine all of their poultry. At the endline of our study, even when 67% of households had a poultry shed, 52% of them still kept at least some of their poultry inside their household dwelling at night, saying that their homes provided better protection from predators, weather, and theft. Other studies have also found that even if households have an outdoor confinement structure, they may still choose to keep some or all of their poultry inside the house at night for similar reasons. One study in rural Ethiopia found that 84% of households had outdoor poultry housing, but 63% of households kept their chickens in the house the previous night; poultry raisers shared that they selected their confinement location to protect chickens from predators and theft or hurting or being hurt by children or larger livestock (Passarelli et al. 2021). Some also mentioned confining their poultry to prevent chickens from wandering off and destroying neighbors’ crops or land. In rural Western Uganda, 41% of households were observed to have some form of poultry housing (a fenced in area, walled enclosure with roof that fully contain poultry, or walled enclosure with roof that poultry can enter and exit freely, all presumably outside, or basket) (Lowe et al. 2022). However, 85% of these households reported keeping at least some poultry inside the house at night, and among households that had no enclosures, 77% kept poultry inside the house at night. In our study, the proportion of households that reported recent predation of chickens was 18% at endline. To address this, sheds could better protect against predators, cold, and theft through stronger materials / smaller openings, improved insulation, and a lock. Behavior change communication may also be required to convince households that these improved sheds would address their concerns.

### Impact of subsidy on shed construction

Our study suggests that subsidies may not be necessary for rural Bangladeshi households to construct poultry sheds. While over >80% of households that received a 23 USD subsidy constructed an improved shed, one-third of households that did not receive a subsidy also constructed an improved shed during the 3-month study period. One reason that more non-subsidy households did not construct sheds during the study may be that the study duration was too short for them to save up sufficient funds or accumulate enough leftover material from other building projects (such as replacing the roof, constructing a shed, or repairing a latrine) to construct the shed. Studies that follow-up with households 6 or 12 months after the intervention may be better positioned to assess the impact of the intervention on households that do not receive a subsidy. Another issue to consider is the availability of trained carpenters who can build sheds. In our study, we found that households used both study-trained carpenters and carpenters not trained by our study. Even in 38% of the households the shed was built by a household member or someone who lives in the same compound. This suggests that finding someone to build the shed may not be difficult in these settings.

Subsidies, when appropriately and accurately targeted and implemented, have the potential to increase equity in sanitation access and use. Past attempts to subsidize sanitation hardware have distorted sanitation markets and created perverse incentives among beneficiaries while primarily benefitting wealthier households and failing to create sustained hardware use (Jenkins and Sugden 2006; Robinson and Gnilo 2016). In addition, the expectation of a subsidy was noted as a constraint to unsubsidized latrine construction in 15% of CLTS interventions (Venkataramanan et al. 2018). However, behavior change promotion with a monetary subsidy has been more effective in increasing latrine coverage (or attenuating declines in latrine use) than behavior change alone (Cameron et al. 2021; Guiteras, Levinsohn, and Mobarak 2015; Garn et al. 2017), so identifying how to most cost-effectively implement subsidy programs and evaluating their effect is useful. Some subsidy approaches, such as vouchers and rebates to cover full or partial costs, can be used to deliver subsidies that stimulate both demand and supply, at least in the short term (Cameron et al. 2021; Guiteras, Levinsohn, and Mobarak 2015; Kohlitz and Iyer 2021). While subsidy validity periods can strongly impact subsidy uptake, vouchers can effectively support households in overcoming liquidity constraints (Kohlitz and Iyer 2021). The optimal amount of the subsidy, which minimizes the implementer’s cost and maximizes uptake, is not clear and likely differs for each household, depending on the total cost of the hardware, the household’s financial status, and the household’s willingness to pay (Peletz et al. 2017). Appropriately targeting subsidies is difficult as different classification methods identify different households as “poor” (Pu et al. 2024). Additionally, households that would benefit from financial subsidies may also need other types of support (such as gender and inclusion initiatives) to harness the long-term benefits of sanitation and other programs (Chambers 1983).

#### Poultry feces presence and disposal

Following the intervention, over 80% of households had no uncontained feces observed inside the house compared to 54% at baseline. The presence of feces inside the house was not associated with the confinement of all poultry outside the house at night or with access to a shed. Even in households where poultry were confined at night in a shed outside, they still roamed freely during the day. The intervention also had no impact on the presence of uncontained feces on the veranda or in the courtyard, suggesting that improvement of conditions inside the house was associated with the recommendation to pick up feces immediately after they are seen. Since this task requires substantial attention and time from the primary poultry raiser, it makes sense that she might choose to focus her efforts only on the inside of the house.

The studies in Ethiopia and Uganda suggest that poultry housing is associated with a lower prevalence and/or count of (presumably, uncontained) feces inside the house or outside in domestic spaces, but details on where feces were observed are insufficient to identify why this difference may have been observed. In Ethiopia, enclosed poultry housing was associated with 1.83 times the odds of having no feces observed outside the enclosure; poultry housing that was 1 to <4 meters from the house was also associated with 1.56 times the odds of no observed feces (Passarelli et al. 2021). In Uganda, 50% of households with an enclosure had no feces observed outside the enclosure compared to 39% of households that had no enclosure (Lowe et al. 2022). Conversely, of the 84% of households that had free-roaming poultry, 62% had observed poultry feces compared to 29% of households that had no free-roaming poultry; it is unclear if these differences are statistically significant.

We encouraged primary poultry raisers to thoroughly scrape animal feces off the ground using a garden hoe with the hope that this would reduce fecal contamination of the underlying and nearby soil. We did not recommend the use of a modified hoe developed specifically for the purposes of animal feces removal (Sultana et al. 2013), but even this modified hoe had low uptake when implemented as part of a multi-component water, sanitation, and hygiene intervention in rural Bangladesh (Parvez et al. 2018).

The percentage of households that had a specific pit for disposing of poultry feces away from the children’s reach increased 44 percentage points from baseline to endline. If used consistently, disposing of feces in such a pit may reduce children’s exposure to poultry feces. However, heavy monsoon rains could flood a feces disposal pit, compost pile, or trash pile or spread fecal across the domestic environment. Fecal matter leaking out of or overflowing from disposal locations into the domestic environment could explain the increased rates of environmental contamination and diarrhea observed following heavy rains (Niven et al. 2023; Nguyen et al. 2024; Mertens et al. 2024). On-site feces containment approaches that are resilient to the weather and climate-sensitive include elevated pit latrines or septic tanks, latrines, twin pit latrines with urine diversion, composting toilets, urine-diverting dry toilets, elevated movable plastic drums, constructed wetland systems, and conventional flush toilets with a biogas system (Borges Pedro et al. 2020). However, these would not allow for beneficial uses of poultry feces, such as fertilizer and fish feed.

### Potential for health impact

While we did not collect data on the prevalence or incidence of diarrhea or other health outcomes, we hypothesize that children had reduced exposure to poultry feces and associated *Campylobacter*. While the intervention did not change the percentage of households that had uncontained feces in the courtyard or veranda, it substantially increased the percentage of households that had no uncontained feces inside the household dwelling. Given that young children in rural Bangladesh are inside about half of their waking hours (Kwong et al. 2016), this reduction in daytime (and nighttime) indoor exposure could result in long-term health improvements if the behavior change was sustained. However, it is also possible that increased confinement had no effect on child health. In a randomized controlled trial in Ethiopia that distributed genetically improved chickens and basic husbandry guidance, households in intervention villages were more likely than households in control villages to have a chicken coop, have a coop separated from the house, and have a coop where chickens were confined at both 9 and 18 months after the intervention. However, the intervention had no impact on the 2-week prevalence of diarrhea, vomiting, fever, or anthropometry at endline (Passarelli et al. 2020). Similarly, in a study conducted in Bangladesh, a homestead food production program with poultry and food hygiene intervention improved ownership of poultry sheds and post intervention among poultry owners, 75% used the shed for poultry, but the intervention had no impact on 1-week diarrhea prevalence (Lambrecht, Waid, et al. 2023; Lambrecht, Müller-Hauser, et al. 2023). Additionally, it is possible that corralling the chickens contributed to worse health outcomes despite the reduced presence of feces indoors because children could have been exposed to a high density of confined feces if they entered the shed. While the average distance from the house to the shed was 15 steps (approximately 10 m) in our study, 36% of respondents reported observing their <5-year-old children entering the shed at least once in the past month. In a randomized controlled trial in peri-urban Peru, households corralling chickens doubled the incidence of *Campylobacter-*associated diarrhea and in households with >20 chickens, the incidence increased by seven-fold (Oberhelman et al. 2006). The authors speculate these adverse health outcomes were because sheds were built on household verandas, making it easy for children to frequently enter the shed, and household members that cleaned the sheds may not have adequately washed their hands afterwards given water scarcity in the community. Given the range of these findings, further research on the impact of poultry housing variations on children’s exposure to *Campylobacter* and diarrheal outcomes is needed to establish the contexts in which corralling is most beneficial and to determine factors affecting the sustainability of behavior change and outdoor coops.

### Limitations

A primary limitation of this pilot study is that it did not have a control group. As such, the trends we observed could have been due to changes over time rather than the intervention. However, given that there were only 3 months between intervention and follow-up, we do not expect to see such large prevalence differences due to time trends alone. While we conducted spot-checks to gather data on the presence of poultry confinement structures, soap and water at handwashing stations, and feces disposal locations, we relied on self-reported handwashing and feces disposal behaviors, and past studies have shown that people tend to over-report behaviors that correspond to the prescriptive norm (Contzen, De Pasquale, and Mosler 2015). Following the baseline survey and prior to initiation of the intervention, Bangladesh imposed nationwide lockdowns due to COVID-19; these lockdowns produced considerable economic hardship (Shahadat, Siddiquee, and Faruk 2020). An even greater percentage of households may construct improved poultry sheds in non-pandemic contexts. In addition, budget constraints limited both the sample size and duration of follow-up, constraining our ability to infer causal mechanisms and assess sustainability.

## Conclusion

The Neighborhood-based Environmental Assessment and Planning (NEAP) intervention successfully encouraged poultry-raising households to construct poultry sheds, confine poultry outside of the household dwelling at night, limit poultry feces from indoor living spaces, and wash hands at key times. Both households that did and did not receive a monetary subsidy constructed improved sheds. While almost all courtyards still had uncontained feces from poultry scavenging for food during the day, children’s overall exposure to poultry feces may have been reduced by the cleaner indoor environment and increased handwashing with soap. Future research could assess if this intervention and approach is able to create sustained use of poultry night sheds, handwashing at key times, and cleaning of indoor spaces. If so, further work could assess the impact of the intervention on clinical and subclinical *Campylobacter* infections and linear growth among young children.

## Data Availability

The data that support the findings of this study can be made available upon reasonable request and approval by icddr,b.

## Acknowledgments

The authors thank all the study participants for their cooperation during the study. The authors acknowledge the contribution of the community hygiene promoters during the intervention delivery and enumerators during the baseline and end line surveys. icddr,b is grateful to the Governments of Bangladesh and Canada for providing core support.

This research was supported by USAID funding through WASHPaLS (USAID contract AID-OAA-I-14-00068/AID-OAA-TO-16-0016). This study was made possible by the support of the American People through the United States Agency for International Development (USAID). The contents of this manuscript are the sole responsibility of authors and do not necessarily reflect the views of USAID or the United States Government. The funders played no part in study design, data analysis, or publication of peer-reviewed results. The funder did require a final report and some of the content of this manuscript is similar to that presented in the final report (USAID and International Centre for Diarrhoeal Disease Research, Bangladesh 2021); all content has been used with permission.

